# Discontinuation of long-acting reversible contraceptive methods and associated factors among reproductive-age women in Shashemane town, Oromia, Ethiopia

**DOI:** 10.1101/2023.07.25.23293161

**Authors:** Fikru Letose, Alemtsehay Tusa, Degemu Sahlu, Yohannis Miherite

## Abstract

**Background:** The early termination of long-acting reversible contraceptives raises issues for the healthcare system and has the potential to affect public health. Long-acting reversible contraception is now more widely available and used, although a sizable percentage of women still stop using it. Therefore, this study aims to assess factors associated with the discontinuation of the long-acting reversible contraceptive method among female users of health facilities in Shashemane Town.

**Methods:** A facility-based cross-sectional study was done in Shashemane town among 410 study participants. The study participants were selected by using a systematic random sampling method. Data were collected by using structured interviewer-administered questionnaires and entered into epi-data version 4.6.0.2 and exported to SPSS version 25 for analysis. Bivariate and multivariable logistic regressions were used to examine the association between independent variables and discontinuation of the long-acting reversible contraceptives. The results were presented using the Odds Ratio at 95% CI. P < 0.05 ware used to dictate statistical significance.

**Result:** The overall prevalence of women who removed the long-acting reversible contraceptive method before the due date was 57.2%. Factors such as having an occupation as a housewife, desire to become pregnant, unwarned side effects, effectiveness, and dissatisfaction with the service provided were positively associated with discontinuation of the contraception.

**Conclusion and Recommendation:** The prevalence of the discontinuation of reversible long-acting contraceptives was high. Pre-insertion effective counseling about the benefits, follow-up care and management for side effects, and client reassurance are recommended.

## Introduction

An unintended pregnancy is defined as a pregnancy that occurred when no children or no more children were desired (Unwanted) or the pregnancy occurred earlier than desired (Mistimed)[1]. It is associated with an increased risk of problems for the mother and baby. According to the World Health Organization (WHO), 74 million women in low- and middle-income countries (LMIC) become pregnant unintentionally each year. This results in 47,000 maternal fatalities and 25 million unsafe abortions each year. This leads to 25 million unsafe abortions and 47,000 maternal deaths every year[2].

One of the most affordable health promotions to lower mother and child mortality by about 30% and 10%, respectively, is the use of modern contraceptive techniques. [3]. Therefore, by ensuring that every pregnancy is intended and every birth is safe, the vast majority of maternal and infant deaths can be avoided.[4]. Long-acting reversible contraceptives (LARCs) are a type of modern birth control method that prevents unintended conception for a long time without forcing the client to exert additional effort for at least three years of continuous use.[5]. Intra-Uterine Copper Devices (IUCDs), the levonorgestrel (LNG)-releasing intrauterine system, and LNG-releasing and etonogestrel (ENG)-releasing subdermal implants are categorized as LARC methods [6–8]. Long-acting reversible contraceptive discontinuation is the removal from using and ceasing it or switching to other methods for any reason before completion of the duration(1).

Despite the improvement in the availability and utilization of long-acting reversible contraception, early discontinuation is becoming a major problem[9]. In low-income countries (LICs), a systematic review report revealed that discontinuation of LARCs within one year of insertion was 20%[10]. In a study conducted on 21 LICs, LARCs comprise**d** only 10% or less, of which 9% of implant users and 15% of IUCD users discontinued the method within the first year of use[11]. Besides, 13.1%, 26.3%, and 36.7% of IUD users discontinued at the first 12, 24, and 36 months, respectively[12]. The Ethiopian Demographic Health Survey (EDHS) revealed that the overall discontinuation of contraceptives at 12 months was 35%, of which 13% were IUD users and 11% were implant users[13]. Other studies conducted in Ethiopia showed that the prevalence of discontinuation of LARCs use was 20%([14], 36.94%([15], 50%[16], and 66.3%[17].

Contraceptive discontinuation may be associated with external factors, individual and/or partner factors [18–20]. The quality of family planning services and obstetric factors like past abortion history, number of living children, desire for more children, preferred family size, sex preference, irregular vaginal bleeding, lower abdominal pain, and abnormal vaginal discharge were linked to LARCs discontinuation, according to a small number of studies[14, 21–26]. Socio-demographic variables such as age, marital status, educational status, and religious concern were found to influence LARC discontinuation. Regarding method-related variables, studies revealed that side effects[11, 14], counseling, satisfaction with the service given, and information about family planning were factors associated with the discontinuation of LARCs[17, 21–23].

In Ethiopia, providing LARCs is a free but highly effective method of preventing unintended pregnancies [27]. Discontinuation while still in need is particularly problematic for women which results in 15% to 20% times at risk of unwanted pregnancy for LARC users three months after discontinuation[11]. Long-acting reversible contraceptives when discontinued to increased fertility rate, unintended pregnancy, and associated complications [11, 28]. The high LARCs discontinuation coupled with low uptake results in a big challenge to achieve the targeted contraceptive prevalence in Ethiopia[29].

Depending on cultural differences and time elapse, the causes for the termination of LARCs may be extremely contextual. Additionally, reducing the discontinuance of LARCs is a great way to prevent or at least minimize unintended pregnancy[30]. In countries like Ethiopia with high fertility and unmet need for family planning[13], it is important to understand how long women continue to use LARCs and what factors are associated with its discontinuation helps to improve the reproductive health of women. However, limited information regarding methods discontinuation from using LARCs and associated factors in the study area. Therefore, the main aim of this study was to determine the discontinuation of LARCs and associated factors among reproductive-age women in Shashemane town, Oromia, Ethiopia.

## Method and materials

### Study area and period

The study was conducted in Shashemene town among women 15-49 years of age. Shashemene town is the capital city of the West Arise zone in the Oromia Region which is 250 Km far from Addis Ababa. The total population of the town is estimated to be 286,447 of which 49.5% are females and 34.7% are reproductive age group women. Currently, the town has two public hospitals, five health centers, two Nongovernmental organizations (NGOs) clinics, 1 private hospital, more than 50 private clinics, and pharmacies are providing health services. From these health facilities, all hospitals, health centers, private hospital, NGO clinics, and nine private clinics provide LARCs services. The study was conducted from May 16 to June 16, 2022.

### Study design

The institution-based cross-sectional study design was applied.

## Populations

### Source population

All reproductive-age women who were using LARCs in the health facilities of Shashemene Town.

### Study population

All randomly selected reproductive-age women who were using LARCs and come to selected health facilities of Shashemene Town with contraceptive-related issues during the study period.

### Inclusion criteria and exclusion criteria

#### Inclusion criteria

All reproductive-age women who were using LARCs and come to the selected health facilities for any issue concerning the method before completion of duration; like removal, side effects, and follow-up during the actual data collection period. Those women who lived in the study area at least for six months were included in the study.

#### Exclusion criteria

All women who have used LARCs outside of Shashemane town and come to the study area for the removal service. Those women who come to change Implanon to Jadelles, Jadelles to Implanon, and implants to IUCD or vice versa were excluded from the study.

### Sample size determination

The quired sample size was calculated by using the population proportion formula equals =

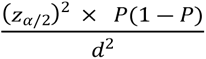

Where *n* is the minimum sample size, *p*=39% prevalence of Implanon discontinuation in Ambo town[31], z=1.96 (95% CI), and d=0.05 margin of error. Applying the formula by substituting these values into the equation gives 373. Ten percent was added for the non-response rate and the final Sample size become 410.

### Sampling Technique

All public health facilities and non-governmental specialty clinics in Shashemene town which provides LARC services were included in the study. Namely, they are two public Hospitals (Referral Hospital and Melk oda G/Hospital), five public health centers (Abosto, Awasho, Bulchana, Dida Boke, and Arada,) and two non-governmental specialty clinics (Family Guidance Association and Marie stops International-Shashemene). Private health facilities were not included in this study because they were not providing full LARC services (insertion and removal). To proportionally allocate the calculated sample size, average client flow for 6 months (December 2021 to April 2022) at each health institution was considered before data collection. The information obtained from the six-month enrollment record of the family planning registration book indicated that a total of 4016 women booked for LARCs at public and non-governmental specialty clinics. The average 6-month client flow identified from the family planning registration book at Referral Hospital, Melk oda General Hospital, Abosto HC, Awasho HC, Bulchana HC, Dida Boke HC, Arada HC, Family Guidance Association and Marie stops International-Shashemene were 452, 419, 403, 410, 354, 349, 322, 618, and 689 respectively. The calculated sample size was proportionally allocated to each facility. Then, the study participants were identified by systematic random sampling method. Data collectors approached and recruited LARC users who had come to the selected clinics for contraceptive-related reasons.

Therefore, every other woman was included in the sample until the total sample size for this study was obtained. The procedure was continued throughout the data collection period until the required sample size was achieved (**Figure 1**).

**Figure 1:**
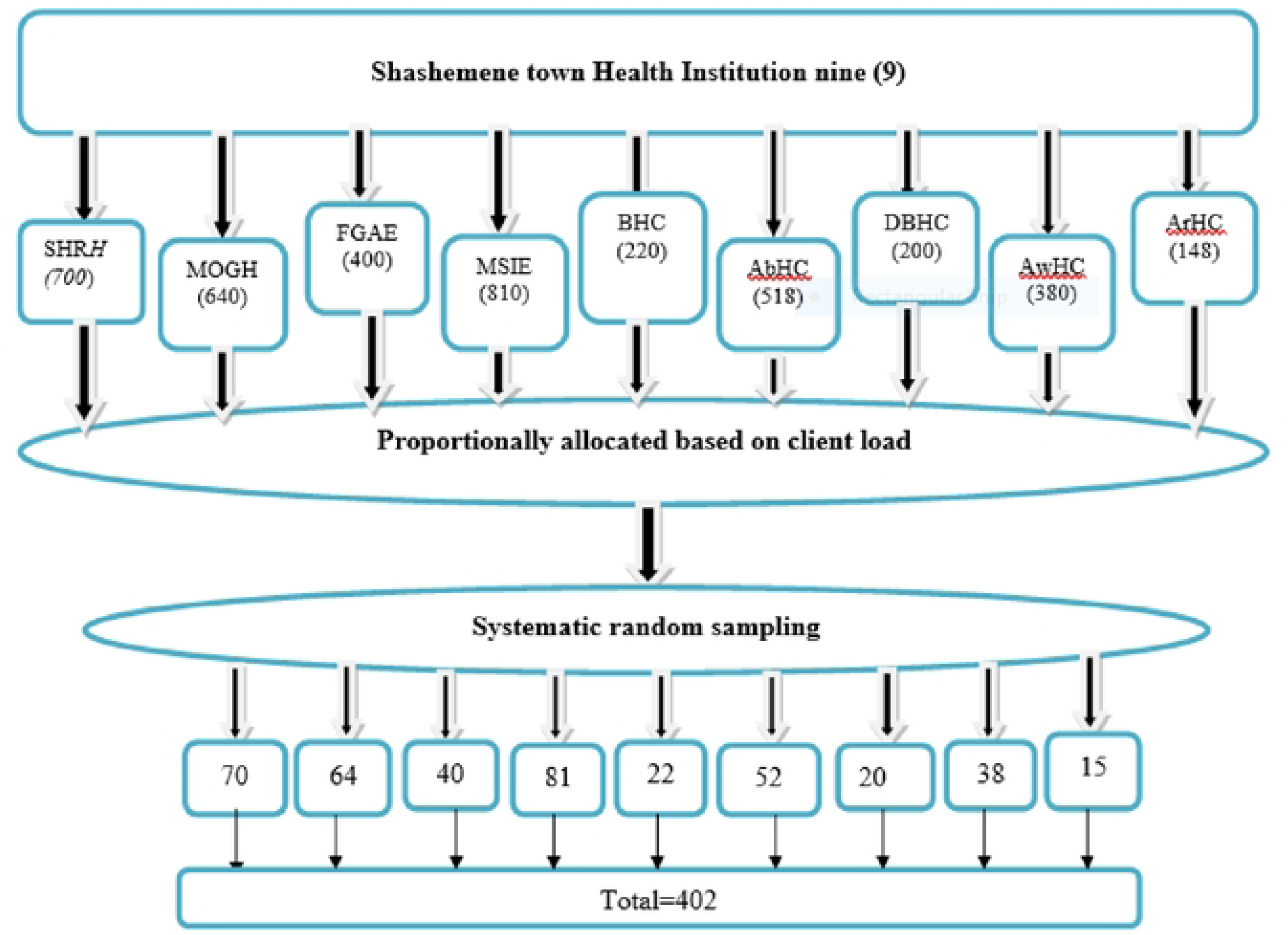
Six months’ enrollment in Heath institutions of Shashemene town administration in two public hospitats, five HCs and two NGO clinics from family planning registration book, for sample selection (December - June/2022).

### Data Collection Tool and procedure

The data were collected using structured questionnaires through the face-to-face interview which is developed after different literature related to the topic under study are reviewed. It was prepared in English and translated to Afan Oromo and Amharic for data collection. The questionnaire contains four parts: Socio-demographic characteristics, Contraceptive and counseling-related characteristics, Health system characteristics, and Obstetric related characteristics. Two days of training were given to data collectors and supervisor about the study objective, procedure, and research ethical rules. Informed consent was taken from selected individual women and getting permission for the data collection takes 15-20 minutes.

### Study variables

The dependent variable was the discontinuation long acting reversible contraceptive method and Independent Variables were Socio-demographic characteristics which are age, marital status, religion, occupation, education, and family income; Health system related characteristics, Health extension workers’ availability and Time taken to reach to health facilities; Obstetrics related characteristics including Parity, Living children, History of abortion, Desire of pregnancy and Time to get pregnancy; Contraceptive and counseling related characteristics Information about contraceptive like History of family planning before used, Type of contraceptive used before LARCs, Responsible body for choosing LARCs, Counseling and Satisfaction with service given.

### Data Quality assurances

To assure the quality of data, the questionnaire was pre-tested during May 9-11/2022 at 01 kebele (Hage) health center which was found outside of the actual study area. Five percent of the calculated sample of women were enrolled to check the validity and reliability of the questionnaire. After pre-testing the questionnaire, revisions and amendments were made accordingly. The training was given to the data collectors on how to interview and check the questionnaire for completeness. The supervisors and principal investigator checked and revised the completeness of the questionnaire and offer necessary feedback to data collectors.

### Data management and analysis

The collected data was entered using Epi-Data version 4.6 and exported to SPSS 23 software for cleaning, recording, categorizing, and analyzing. A bivariate analysis was done to see the association between independent and outcome variables. Those variables with a P-value of 0.2 during the bivariate analysis were included in the multiple logistic regression analysis to assess the relative effect of confounding variables. Since the outcome variable is categorical, the adjusted odds ratio was calculated using a multiple logistic regression model. After multivariate analysis had been done, the adjusted odds ratio (OR) was used to measure the strength of the association between the dependent variable and the independent variable, while the 95% CI and P-value were used to assess whether the association was significant.

### Ethical consideration

An ethical assurance letter was received from Salale University (SLU) College of Health Sciences, Department of Public Health. It was presented to the Shashemene town health office to grant official permission to undertake research activities in the selected health facility. Written informed consent was obtained from each participant after the investigator had explained the nature, purpose, and procedure of the study. The signature of each participant was used on the willingness confirmation. The entire study’s participants were informed that data was kept private and confidential and used only for research purposes. The participants were also assured that they have the right to refuse or withdraw if they are not comfortable at any time.

## Results

### Socio-demographic Characteristics

A total of 402 women responded to the questionnaire forming, a response rate of 98%. The mean age of the respondents was 26 years (± 4.6). About 241(60.0%) were Oromo by Ethnicity, and 150(37.3%) were Muslims by religion. Most of the respondents 268(91.5%) were married. Two hundred fifty-three (62.9%) respondents attended secondary education. Regarding occupation, the majority of the respondents 230(57.2%) were housewives. More of the respondents have a monthly income of 2000-3000 ETB per month (**Table-1**)

**Table 1:**
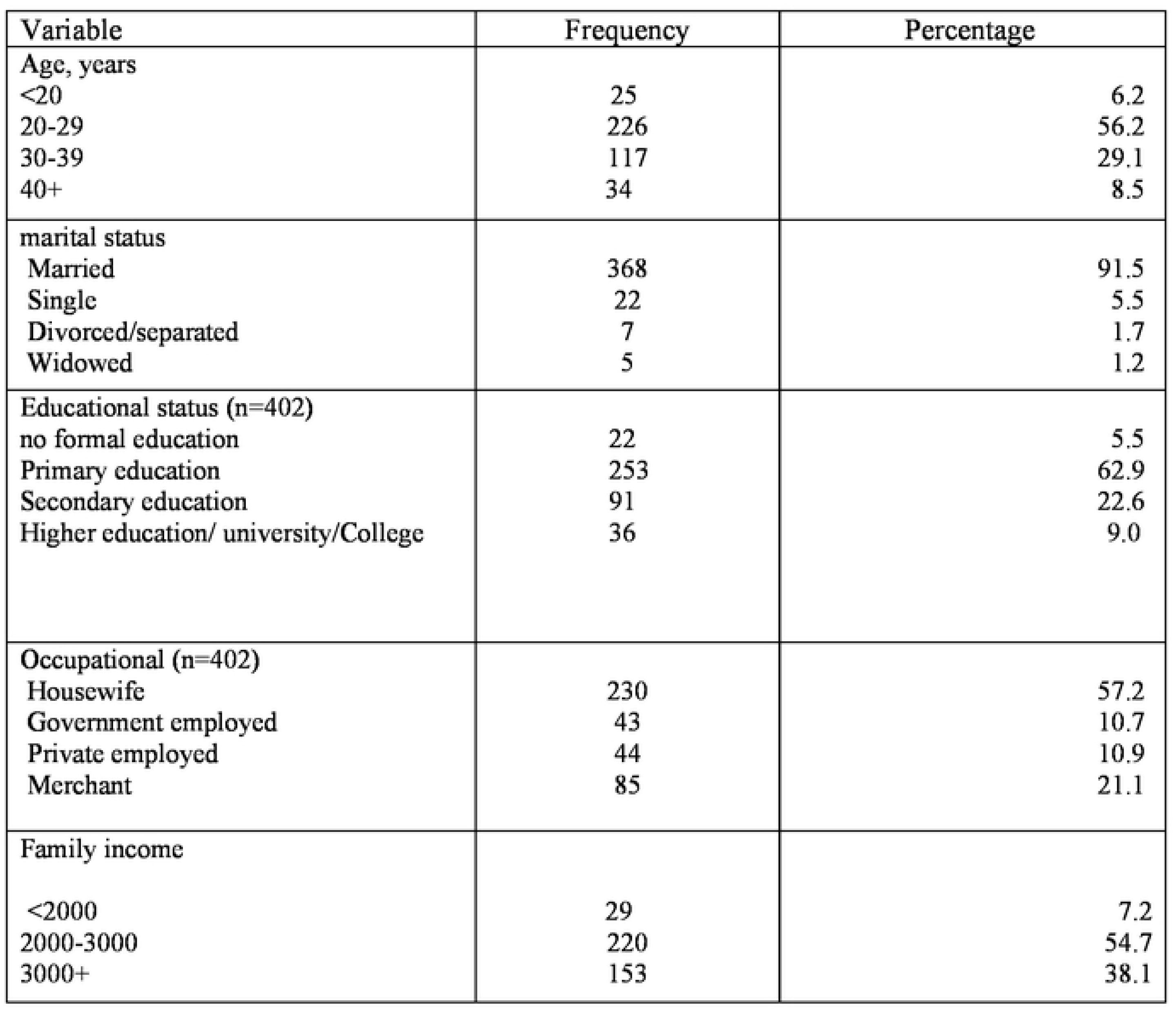
Socio-demographic characteristics of long-acting reversible contraceptive users in health facilities of Shashemene town, Southern Ethiopia, 2022(n=402).

### Health system-related characteristics

The majority of women 342(85.1%) have access to health extension workers in their catchment and more than half of the women 252(62.7%) live 30-60 minutes walking distance from the health facilities.

### Obstetric-Related Characteristics

Among the women who ever used LARCs, only three (1%) have no history of live birth. Besides, 9 (2.2%) have no child, 122(30.3%) have a history of abortion, and 177(44%) desire to become pregnant (**Table- 2.**)

**Table 2:** Obstetric history of women who ever used LARC 2021/2022, in Shashemene town, Oromia, South Ethiopia, 2022(n=402).

| Variable | Frequency | Percentage |
| --- | --- | --- |
| Parity |  |  |
| 0 | 3 | 0.7 |
| 1-2 | 137 | 34.1 |
| 3-4 | 203 | 50.5 |
| 5+ | 59 | 14.7 |
| Living children (n=198) |  |  |
| 0 | 9 | 2.2 |
| 1-2 | 139 | 34.6 |
| 3-4 | 198 | 49.3 |
| 5+ | 56 | 13.9 |
| History of abortion |  |  |
| Yes | 122 | 30.3 |
| No | 280 | 69.7 |
| Desire of pregnancy |  |  |
| Yes | 177 | 44.0 |
| No | 225 | 56.0 |

### Contraceptive and counseling-related characteristics

From the total number of women interviewed, about three-fourths (74.4%) used other contraceptive methods before LARCs. For instance, of these women, about 150(50.3%) had been using injectables. Regarding method selection, nearly two-thirds (64.7%) of women decided to use LARCs by themselves. Among the type of LARCs, 220(54.7%) women used Implanon. Among the total study participants, those women counseled about the benefit, side effects, and effectiveness of LARCs were 199 (49.5%), 203 (50.5%), and 227(56.5%) respectively. One hundred ninety-five (48.5%) of respondents mentioned having been satisfied with the service given before the insertion (**Table-3**).

**Table 3:** Contraceptive and counseling-related characteristics of women who ever used LARCs in 2021/2022 in Shashemene town, Oromia, Southern Ethiopia,2022(n=402).

| Variable | Frequency | Percentage |
| --- | --- | --- |
| Contraceptive use before LARCs |  |  |
| Yes | 299 | 74.4 |
| No | 103 | 25.6 |
| Type of contraceptive used before LARCs |  |  |
| Injection | 150 | 50.2 |
| OCP | 93 | 31.1 |
| OCP nad Injection | 45 | 15.2 |
| Others | 10 | 3.5 |
| Method selection |  |  |
| My self | 260 | 64.7 |
| My husband | 12 | 3.0 |
| Health professional | 83 | 20.6 |
| Others (Health Extension workers, Neighbor...) | 47 | 11.7 |
| Type of LARC used |  |  |
| Implanon | 220 | 54.7 |
| Jaddles | 110 | 27.4 |
| IUD | 72 | 17.9 |
| Counseled about benefit of LARC |  |  |
| Yes | 199 | 49.5 |
| No | 203 | 50.5 |
| Counseled about side effect |  |  |
| Yes | 203 | 50.5 |
|  | 199 | 49.5 |
| No |  |  |
| Counseled about effectiveness |  |  |
| Yes | 227 | 56.5 |
| No | 175 | 43.5 |
| Information participants got during counselling (Satisfied) |  |  |
| Yes | 216 | 53.7 |
| No | 186 | 46.3 |

### The magnitude of long-acting reversible contraceptive discontinuation

The study revealed that among the study participants who ever used LARCs, 18.4%, 57.2%, and 81% of women discontinued the service at 12, 24, and 36 months, respectively. Besides, 38.8% and 23.8% of women discontinued it at 12-24 and 24-36 months, respectively. Among the study participants, only 76(19%) of women used LARCs for at least 36 months duration. Regarding the types of LARCs discontinued at 12 months, the proportion of women who discontinued Implanon, Jadelle, and IUCD was 51.4%, 31.1%, and 17.6% respectively **(Figures 2**, **3, and 4**.)

**Figure 2:**
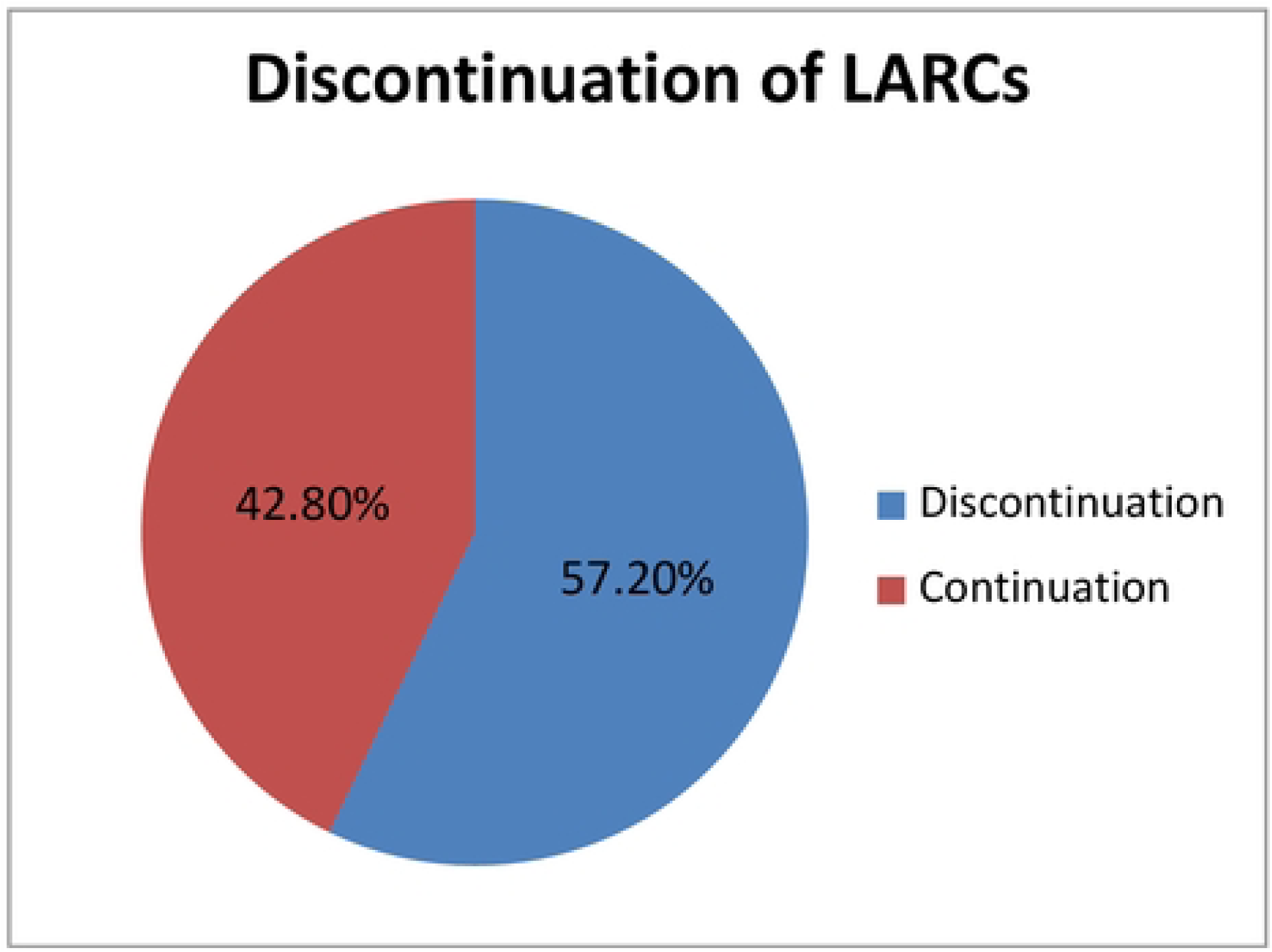
proportion of LARC discontinuation rate among LARC users in Shashemene town, Southern Ethiopia, 2022.

**Figure 3.**
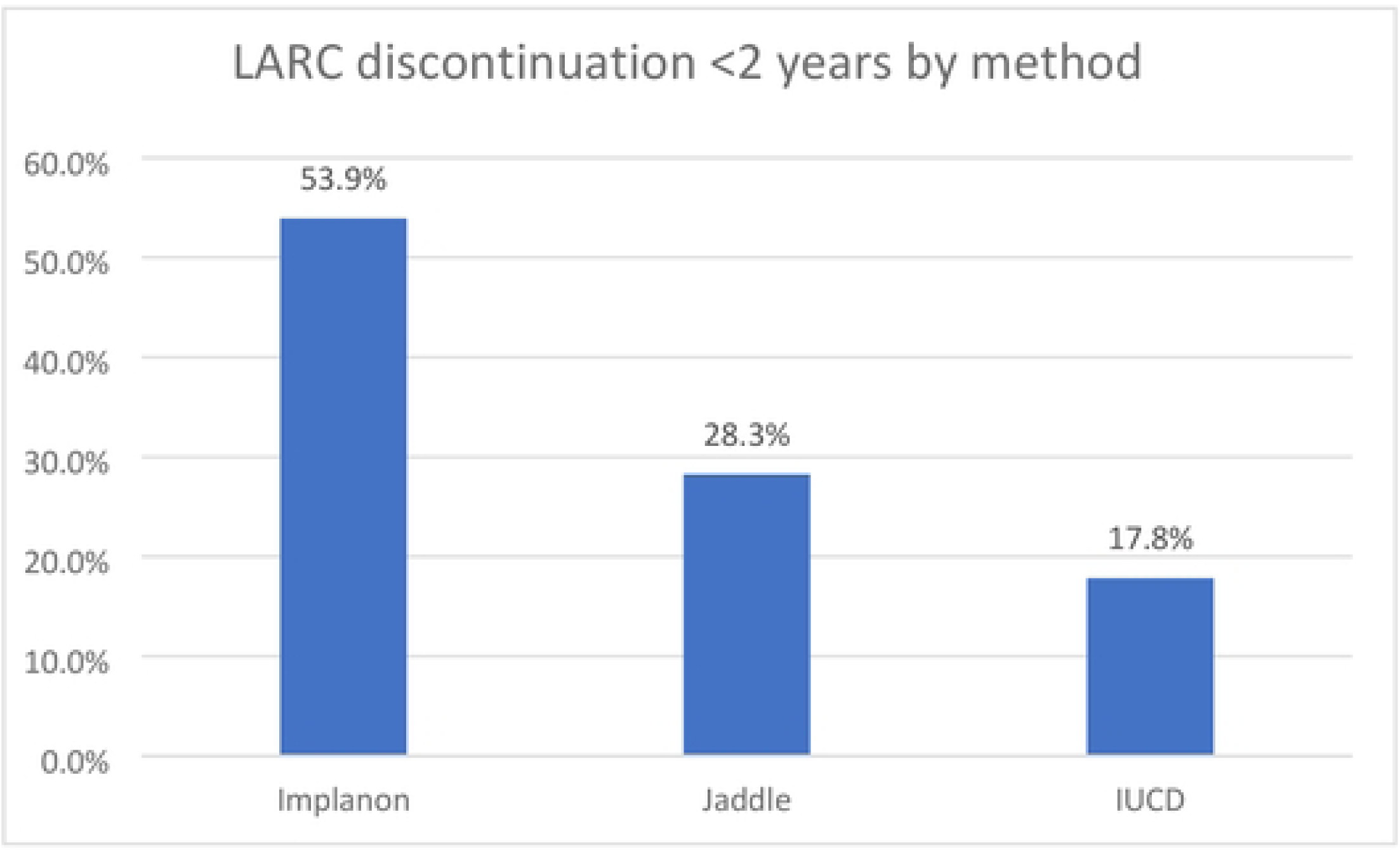
LARC discontinuation <2 years by method.

**Figure 4.**
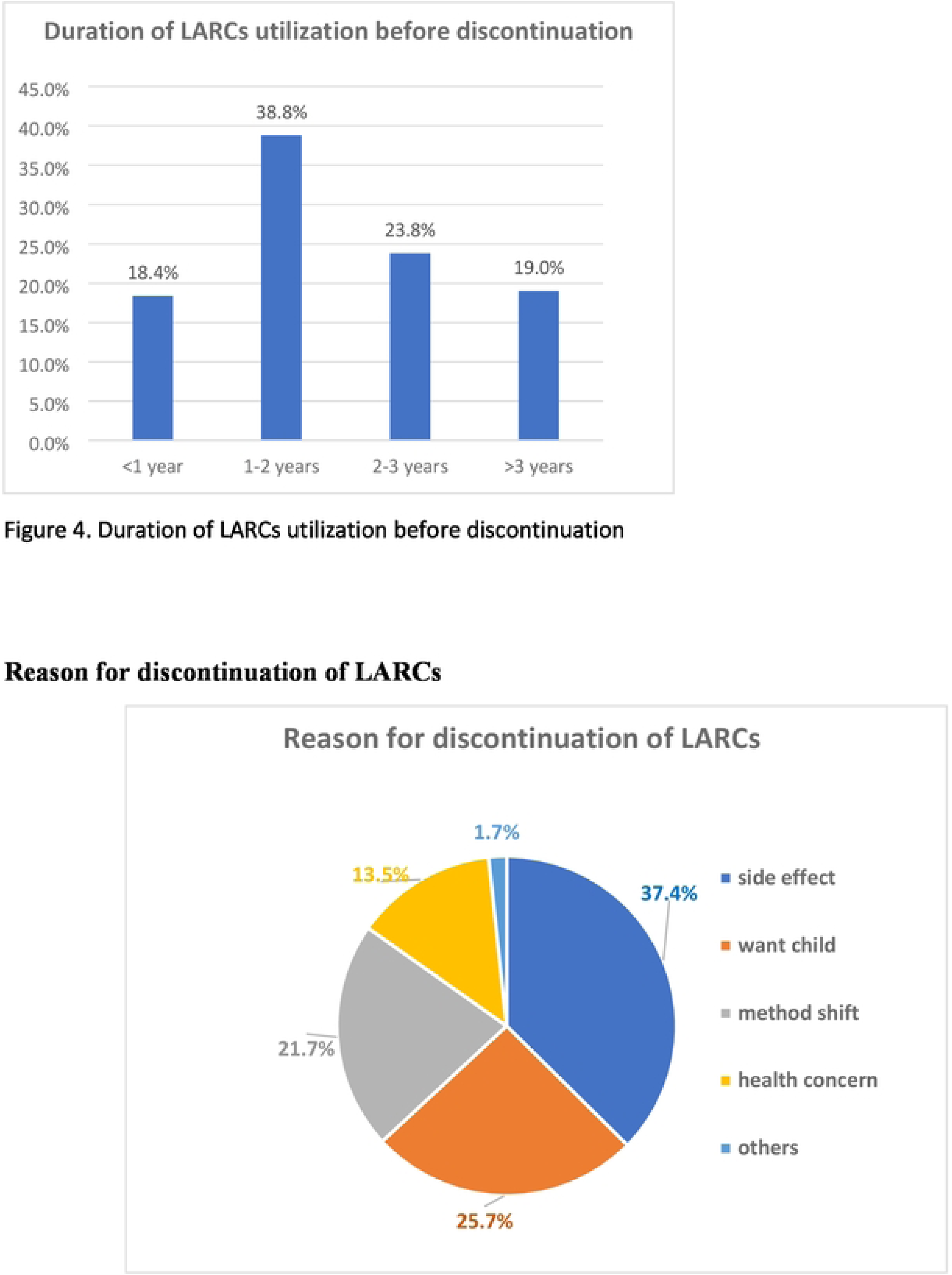
Reason for discontinuation among women who ever used LARC in 2021/2022 in Shashemene town, Southern Ethiopia (n=230).

#### Reason for discontinuation of long-acting reversible contraceptives

More than one-third (37.4%) of women discontinued LARCs within three years of utilization, mainly due to emerging side effects, followed by 84 (25.7%) who desired to have a child **(Figure -5)**. The most common complaints among women who developed side effects were menstrual disruption 58(47.7%), weight gain 24(19.8%), insertion site arm pain 17(14.0%), unusual headache 16(12.8%), infection 6(4.7%) and method inconvenience1(1.2%) **(Figure -5.**)

**Figure 5.**
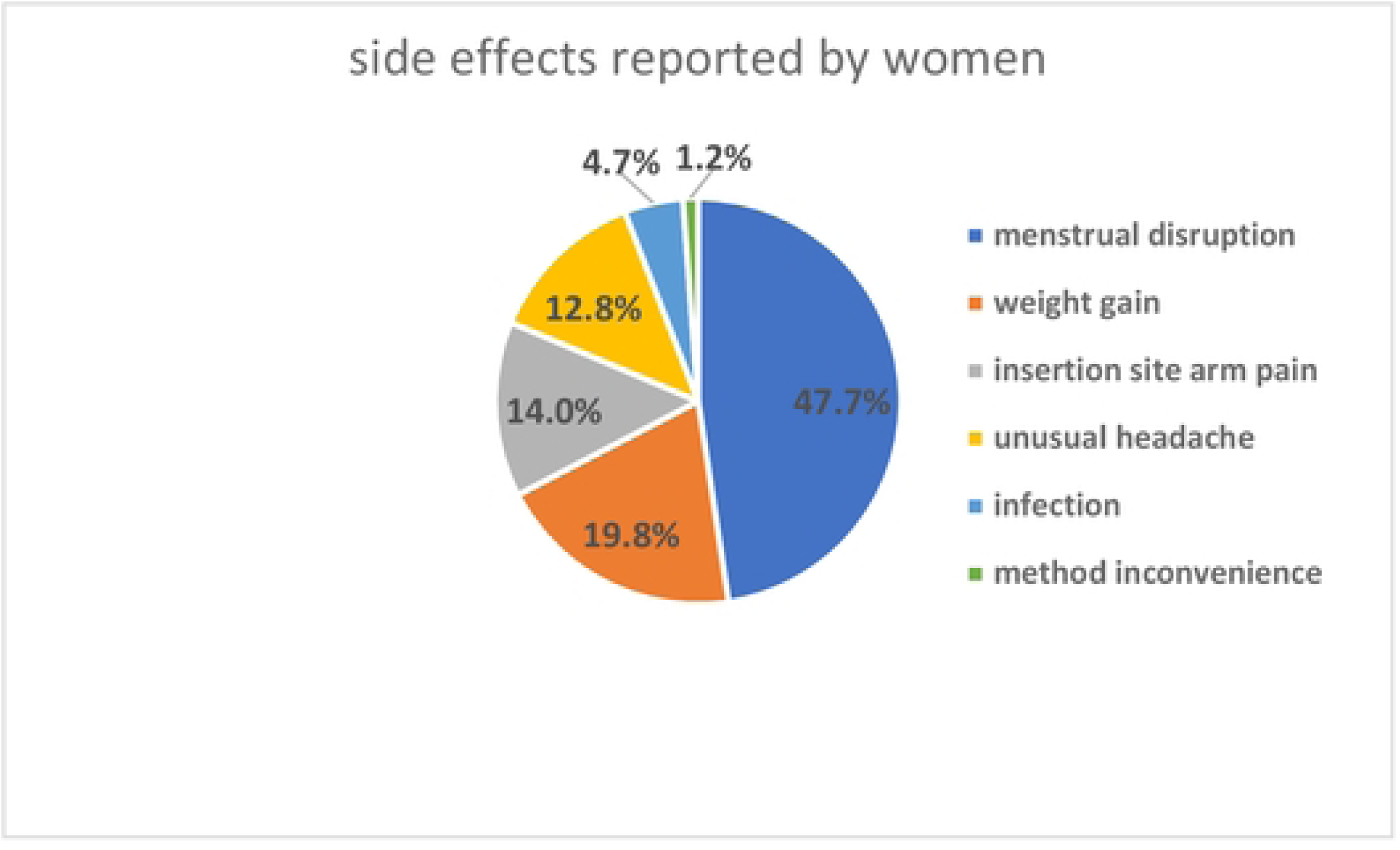
side effects reported by women.

### Factors associated with discontinuation of reversible long-acting contraceptives

From the bivariate logistic regression analysis, factors such as age, educational status, occupational status, desire to become pregnant, counseling about the benefit of LARCs, counseling on the effectiveness of LARCs, and satisfaction with the service given have a *p*-value ≤0.25 and are considered for multivariable logistic regression.

Factors from bivariate logistic regression analysis with a *p*-value ≤0.25 were entered into multivariable logistic regression analysis. With multivariable logistic regression analysis, considering a p-value ≤0.05, the independent predictors of LARCs discontinuation were maternal occupation, desire to be pregnant shortly, lack of counseling about the side effects of LARCs, no counseling on the effectiveness of LARCs, and not being satisfied with LARCs service provision **(Table 4.**)

**Table 4:** Bivariate and Multivariate analysis of factors associated with discontinuation of long-acting versible contraceptives in Shashemene town southern Ethiopia, 2022 (n=402).

| Variables | Discontinuation of LARCs |  | COR (95%CI) | P-value | AOR (95%CI) | P-value |
| --- | --- | --- | --- | --- | --- | --- |
|  | Yes% | No% |  |  |  |  |
| Age |  |  |  |  |  |  |
| <20 | 13(52.0%) | 12(48.0%) | 1 |  |  |  |
| 20-29 | 162(71.7%) | 64(28.3%) | 0.43(0.18- 0.98) | 0.047 | 0.59(0.17 - 1.98) | 0.394 |
| 30-39 | 41(35.0%) | 76(65.0%) | 2.00(0.84 -4.80) | 0.117 | 2.34(0.68 - 8.02) | 0.175 |
| 40+ | 14(41.2%) | 20(58.8%) | 1.54(0.54 -4.38) | 0.411 | 5.53(0.26 –24.16) | 0.073 |
| Educational status |  |  |  |  |  |  |
| no formal education | 10(45.5%) | 12(54.5%) | 1 |  | 1 |  |
| Primary education | 191(75.5%) | 62(24.5%) | 0.27(0.11- 0.65) | 0.004 | 0.47(0.14 - 1.56) | 0.220 |
| Secondary education | 24(26.4%) | 67(73.6%) | 2.33(0.89 -6.08) | 0.085 | 4.82(0.24 - 18.67) | 0.093 |
| Higher education | 5(13.9%) | 31(86.1%) | 5.17(1.46-18.27) | 0.011 | 8.24(0.41 - 47.96) | 0.099 |
| Occupational status |  |  |  |  |  |  |
| Housewife | 184(80.0%) | 46(20.0%) | 0.09(0.05 -0.17) | 0.000 | 0.09(0.04 - 0.17) * | 0.000 |
| Government employee | 4(9.3%) | 39(90.7%) | 3.83(1.23-11.90) | 0.020 | 0.69(0.15 - 3.09) | 0.630 |
| Private employee | 18(40.9%) | 26(59.1%) | 0.56(0.26 - 1.22) | 0.147 | 0.45(0.16 – 1.27) | 0.135 |
| Merchant | 24(28.2%) | 61(71.8%) | 1 |  | 1 |  |
| Desire of pregnancy |  |  |  |  |  |  |
| Yes | 104(46.2%) | 121(53.8%) | 2.87(1.89 - 4.36) | 0.000 | 3.69(1.90–7.14) * | 0.000 |
| No | 126(71.2%) | 51(28.8%) | 1 |  | 1 |  |
| Counseled about benefit of LARC |  |  |  |  |  |  |
| Yes | 74(37.2%) | 125(62.8%) | 5.60(3.63 –8.66) | 0.000 | 0.79(0.29 – 2.12) | 0.643 |
| No | 156(76.8%) | 47(23.2%) | 1 |  | 1 |  |
| Counseled side effect of LARC |  |  |  |  |  |  |
| Yes | 152(76.4%) | 47(23.6%) | 1 |  | 1 |  |
| No | 78(38.4%) | 125(61.6%) | 5.18(3.36 –7.98) | 0.000 | 2.57(1.10 -6.01) * | 0.029 |
| Counseled effectiveness |  |  |  |  |  |  |
| of LARC |  |  |  |  |  |  |
| Yes | 122(69.7%) | 53(30.3%) | 1 |  | 1 |  |
| No | 108(47.6%) | 119(52.4%) | 2.53(1.67 - 3.83) | 0.000 | 2.77(1.43 - 5.39) * | 0.003 |
| Satisfied with the service given |  |  |  |  |  |  |
| Yes | 162(78.3%) | 127(65.1%) |  |  | 1 |  |
| No | 68(34.9%) | 45(21.7%) | 6.72(4.31-10.46) | 0.000 | 2.83(1.15 -6.95) * | 0.023 |

\*p<0.05, OR- Odds Ratio, AOR- Adjusted Odds Ratio

Among socio-demographic variables, the occupational status of women was found to be associated with the discontinuation of LARCs. Women having the occupation of housewife were 10% times less likely to discontinue LARCs than women having the occupation of merchant (AOR=0.09; CI: 0.04, 0.17). The odds of discontinuing LARCs among women who desire to be pregnant were 3.7 times higher than counterparts (AOR=3.7; CI: 1.90, 7.14). Regarding counseling status of women about the side effects of LARCs, those who were not obtained counseling would be 2.6 times more likely to discontinue LARCs than their counterparts (AOR=2.6; CI: 1.10,6.01). Those Women who were not counseled about the effectiveness of LARCs were 2.8 times more likely to discontinue LARCs than their counterparts (AOR=2.8; CI: 1.43, 5.39). Women who were not satisfied with the service given were 2.8 times more likely to discontinue LARCs than their counterparts (AOR=2.8; CI: 1.15, 6.95) **(Table 4.**)

## Discussions

Nevertheless, 30% of maternal and 10% of infant deaths reduced by LARCs which is one of the most practical and cost-effective method by ensuring that every pregnancy is desired and every birth is safe[3, 4], the overall prevalence of LARCs were found to be 18.4%. Current study finding was consistent with the studies conducted at Hawassa City from Southern Ethiopia of 17.7% [29], Bahir Dar City from Northwest Ethiopia of 17.2% [32] and slightly less than urban public health facilities of Ethiopia[32].

The current study’s percentage of LARCS discontinuation at 36 months was lower than the 65% found in a study conducted in Debre Tabor, Northwest Ethiopia. This variation could be a result of the subjects or the design of the study. Participants from Debre Tabor, in contrast to the current study, included both urban and rural populations. Urban inhabitants might be more aware of the consequences of contraception. Additionally, efforts may have been taken to enhance counseling, particularly for the Women in the current study who had issues with weight gain and menstruation disruption. The current study result was also lower than studies conducted in Ambo 38.2%(26), Dale District 23.4%(27), and Arsi zone 25%(19). This discrepancy might be due to differences in the study design, sample size, duration of the study, lack of pre-insertion counseling, satisfaction with the given service, and sociocultural differences of respondents across the study areas.

Among the types of LARCs discontinued in this study, the prevalence of Implanon (41.8%) and IUCD (15.2%) were found to be less than secondary data analyzed from EDHS of 61% and 45% by 36 months respectively (12). This might be due to the discrepancy in the population structure of the national pattern and variation in the duration of utilization before discontinuation.

The study revealed that the main reason for discontinuation of LARCs was emerging side effects (37.4%), desire to have a child (25.7%), method shift (21.7%), and health concern (13.5%). Menstrual disruption (47.7%) was the main side effect faced among discontinuers. Menstrual irregularity may not cause serious health problems but interferes with daily activity, especially sexual activity with their husband. Discontinuation due to side effects was a result of inadequate counseling and information on possible side effects before and following the insertion of LARCs. Giving adequate counseling and information on the side effects and effectiveness of LARCs likely improves service satisfaction and ensure the continuation of LARCs [13, 16, 29].

Among the independent predictors, the occupational status of women was found to be associated with discontinuation LARCs. Women in the occupation of housewives were 91% more likely to discontinue LARCs than merchants. This may be because women having the occupation of merchant might be more likely to be educated and have access to information about modern family planning from different sources.

The current study also revealed that the odds of discontinuing LARC among women who desired to be pregnant shortly were 3.6 times higher than their counterparts. This is consistent with studies conducted in Kenya[33], Bahir Dar City [34], Debre Markos town[35], and Hawassa City[29]. This might be because the culture of the community influences married women to bring a child to ensure the ability of the women to bear a child. In addition to this, 62.4% of women in the present study were aged 18–29 years. Since the majority of study participants in this study were young, they might intend to have more children and discontinue the contraceptives.

The present study showed that women who were not counseled about the side effects of the LARCs were 2.6 times more likely to discontinue LARCs compared to their counterparts which are consistent with the study done in Ethiopia, systematic review[13], Hawassa[29], and Kersa district[36]. Besides, the current study also revealed that counseling women about the effectiveness of LARCs were negatively associated with LARCs discontinuation. The most cited reason for this effect was effective counseling at insertion time on possible side effects of the method will help the mother to accept the possible side effects and minimize removal of the method early. In addition to this, the use of appropriate counseling type and timing might also help the women to cope with minor side effects and strengthen the continuation of the method.

According to the study, women who were unsatisfied were 2.8 times more likely to stop using long-acting reversible contraceptives (LARCs) than those who were satisfied. This might be the result of incorrect pre-insertion counselling and a lack of seclusion, which led to frustration and quitting. This finding is consistent with a study conducted in Makelle City, Dale district, Kambata zone in southern Ethiopia, and Vietnam(18,21,27,36). This might be due to women’s lack of interest in the method chosen. Furthermore, the confidentiality of the service, elucidation of the service supplier, communication skills, and other service provisions during the insertion of LARCs may contribute to being discontinuity of long-acting reversible contraceptives. Therefore, providing quality reproductive health services to clients can lead to better satisfaction and use of the services.

### Limitation

The study’s cross-sectional design prevented it from determining the cause-and-effect link. In addition, the possibility of recall bias could not be ruled out, and to study determinant factors, a cross-sectional design study has limitations.

### Conclusion

In this study, the overall discontinuation of LARC among women was high (57.2%). The independent predictors of LARC discontinuation among women were maternal occupation as a housewife, the desire for pregnancy, not counseling on side effects, not counseling on the effectiveness of LARCs, and not being satisfied with the service given.

## Recommendation

Address low community awareness through community-level awareness creation programs and health education to ensure long-acting reversible contraceptive use. Provide pre-insertion counseling on side effects, effectiveness, and benefits. Early management and reassurance can decrease discontinuation and enhance retention. A further comprehensive study of this area that includes health professionals and is community-based is recommended.

### Consent for publication

All authors read the manuscript and have provided their consent to publish

### Availability of data

The data sets used and/or analyzed during the current study are available from the corresponding author upon reasonable request.

## Data Availability

All relevant data are within the manuscript and its Supporting Information files.

## Competing Interest

All authors declare that they have no Competing Interest

## Funding

The funding source for per diem of data collectors and supervisors of this research was Salale University. The funding body has no role in the design of the study and collection, analysis, and interpretation of data and in writing this manuscript.

## Notes

### Competing Interest Statement

The authors have declared no competing interest.

### Funding Statement

The funders had no role in study design, data collection and analysis, decision to publish, or preparation of he manuscript.

### Author Declarations

The institutional ethical review committee of Salale University with the Ethical Approval number of SIU 321/2022

